# The Association of Racial Residential Segregation and Long-Term Outcomes after Out-Of-Hospital Cardiac Arrest Among Medicare Beneficiaries

**DOI:** 10.1101/2022.10.05.22280740

**Authors:** Ethan E Abbott, David G Buckler, Jesse Y Hsu, Benjamin S Abella, Lynne D. Richardson, Brendan G Carr, Alexis M Zebrowski

## Abstract

**Background:** Racial residential segregation in the US is associated with poor health outcomes across multiple chronic conditions including cardiovascular disease. However, the national impact of racial residential segregation on out-of-hospital cardiac arrest (OHCA) outcomes after initial resuscitation remains poorly understood. We sought to characterize the association between measures of racial and economic residential segregation at the ZIP code level and long-term survival after OHCA among Medicare beneficiaries.

**Methods:** In this retrospective cohort study, utilizing Medicare fee-for-service claims data from 2013-2015, our primary predictor was the index of concentration at the extremes (ICE), a measure of racial and economic segregation. The primary outcomes were death at 1 and 3 years. Using random-effects Cox proportional hazards models, including a shared frailty term to account for clustering at the hospital level, we estimated hazard ratios across all three types of ICE measures for each outcome while adjusting for beneficiary demographics, treating hospital characteristics, and index hospital procedures.

**Results:** We identified 29,847 OHCA claims for beneficiaries who survived to discharge after an OHCA. Mean beneficiary age was 75 years (SD 8); 40.1% were female, 80% White and 15.2% Black. Overall crude survival for the cohort was 54% (n=16,129) at 1 year and 40.8% (n= 12,189) at 3 years. In fully adjusted models we found a decreased hazard of death in beneficiaries residing in the most racially and economically privileged ZIP codes (Q5) compared to the least privileged areas (Q1) across all three ICE measures (race: HR:0.84; CI 0.79-0.88, income: HR 0.76; CI 0.73-0.81, race + income: HR 0.78; CI 0.74-0.83)

**Conclusion:** We found a decreased hazard of death for those residing in predominately White and higher income ZIP codes as compared to majority Black and lower income ZIP codes when using validated measures of racial and economic segregation. Future work will need to more closely examine the causal pathways and mechanisms related to disparities in outcomes after OHCA to better understand the impacts of spatial and living environments on long-term outcomes.

## Introduction

Over 400,000 cases of out-of-hospital cardiac arrest (OHCA) occur each year in the U.S with overall low rates of survival to discharge and often with poor functional neurological status.^1-4^ Known racial and ethnic disparities in the incidence and short-term survival following OHCA have been identified across multiple studies,^2, 3, 5-10^ and prior work has established that overall, OHCA survivors are a vulnerable patient population; high mortality and frequent readmissions are noted among those who survive to discharge from their index hospitalitalization.^11, 12^ However, longitudinal outcomes data among OHCA survivors are difficult to ascertain, resulting in an important knowledge gap. As a result, questions remain regarding the magnitude of disparities following initial resuscitation.

Racial residential segregation, the result of longstanding structural and institutional racism in the US, is associated with poor outcomes across multiple chronic health conditions including several cardiovascular diseases.^13-17^ Our prior work examining the association between measures of racial and economic residential segregation and survival to discharge after OHCA demonstrated a lower likelihood of survival to discharge among Medicare beneficiaries residing in predominately Black and lower income ZIP codes as compared to White and higher income ZIP codes.^18^ The mechanisms of residential segregation on health outcomes are complex and multifactorial but are likely to impact quality and access to healthcare, and are also spread across other social domains within each community. OHCA survivors residing in disadvantaged areas likely experience amplified disparities due to the structural and community effects of segregation that affect health and social resources along their continuum of care.

In this study we sought to characterize the association between measures of racial and economic residential segregation at the ZIP code level and long-term survival up to 3 years after OHCA. This was specifically examined among Medicare beneficiaries while accounting for beneficiary demographics, treating hospital characteristics, and procedures performed during index hospitalization.

## Methods

### Study Population

We performed a retrospective observational cohort study, utilizing age-eligible (≥65 years old) Medicare fee-for-service claims data from the Medicare Provider Analysis and Review (MedPAR) and Outpatient Research Identifiable Files (RIF) for January 2013 – December 2015. We abstracted individual patient demographics including race/ethnicity, sex, and age from the Medicare Beneficiary Summary file. Beneficiary date of death was determined from the Vital Status File containing validated dates of death up to June 2019. Therefore, all beneficiaries had at least 3.5 years of follow up data. Claims were excluded if the patient age was <65 years at the start of the study period. Patient-level race was characterized as Black, White, or Other due to the small distribution of beneficiaries with race or ethnicity other than White or Black in the cohort, and the final analysis focused on outcomes between Black and White beneficiaries.

We identified beneficiaries with Emergency Department (ED) treated OHCA, using claims with ICD-9-CM codes 427.5, 427.4, 427.41, 427.42 and mapped ICD-10-CM codes I46, I49.0, I49.01, I49.02 (International Classification of Diseases, Ninth and Tenth Revision, Clinical Modification) as the primary or admitting diagnosis, based on prior approaches utilized for identifying OHCA patients.^19-22^ From this group, we identified beneficiaries who survived to discharge using date of discharge and validated date of death. The primary outcome, survival up to 3 years, was identified using beneficiary death date. Beneficiaries with no documented date of death were censored on 1/1/2019. Interhospital transfer after index OHCA were included in models as a separate covariate.

#### Index of Concentration at the Extremes

Our primary predictor of interest was the validated Index of Concentration at the Extremes (ICE) which measures racial, income and racialized income residential segregation following a specific approach that has been refined and adapted by Krieger and colleagues. ^23, 24^ They demonstrated that measures of racialized economic segregation are more robust at identifying associations with poor health outcomes and avoid potential methodological issues encountered using other approaches.^16, 23-25^

The ICE measure is calculated as the difference between the count of the privileged population and disadvantaged population, divided by the total population. The index is the scaled difference between the population counts of the more privileged group and the less privileged group. Based on previous work using ICE, we calculated three measures including: race, income, and race and income combined^24^ (derived from US Census ACS five-year estimates). US Census Bureau American Community Survey (ACS) five-year estimates were used to calculate ZIP-code tabulation area (ZCTA) -level racial composition and income ICE measures. These measures were then mapped to the claim based on the residential ZIP code on the index OHCA claim. Beneficiary residence was determined from the primary claim present at the index admission. All estimates were calculated at the ZIP code level.^26^ Black and White race were utilized for racial ICE calculations as these two groups represent the extreme ends of racial privilege and deprivation for the US^23^ and were also the most representative among the Medicare datasets. For the ICE measures of income, we defined the lowest income group as <$25,000 and ≥$100,000 for the highest income group, representative of the 20^th^ and 80^th^ percentiles of income for the US.^16^ ZIP code level ICE scores were then binned into quintiles, with Q5 representing the most economically advantaged ZIP codes and Q1 representing the least economically advantaged ZIP codes. For ICE measures of race and income combined, we examined high-income White and low-income Black based on prior work.^16, 24, 25, 27^ This approach functions to represent the extreme ends of racialized economic segregation in the US.

#### Covariates of interest

##### Patient-level

Beneficiary comorbidities were identified from secondary diagnoses present on admission in the MedPAR or outpatient RIF and summarized using the AHRQ Elixhauser Comorbidity index.^28, 29^ Beneficiary age category, sex and race were pulled from the index hospital claim. OHCA length of stay (LOS) was calculated from the first admission date to the final discharge date, including transfers to other short-stay hospital claims. Additionally, we identified cardiac catheterization and implantable cardioverter defibrillator (ICD) placement procedures performed at the first treating hospital.

#### Hospital-level

To address confounding from treating hospitals, we selected characteristics from the American Hospital Association Survey dataset. Hospital characteristics include number of hospital beds, hospital teaching status, availability of medical and cardiac intensive care beds, and hospital ownership (for profit, non-profit, government/non-federal).

### Statistical analysis

Descriptive statistics were calculated using means with standard deviation or medians with inter-quartile ranges, as appropriate for continuous covariates of interest, and frequencies with proportions for nominal variables. To estimate the association of OHCA mortality and ZIP code ICE measures we constructed three models: 1) ICE race, 2) ICE income, and 3) ICE racialized income (income + race). We first estimated survival using Kaplan-Meier methods for each of the ICE measures independently. We then subsequently fitted a random-effects Cox proportional hazards model, including a shared frailty term to account for clustering at the hospital level, to estimate hazard ratios across all three ICE measures. All analyses and graphics were conducted using standard statistical software (R version 4.04, R Core Team: Vienna, Austria). This study was completed in accordance with the STROBE guidelines.^30^ The study was reviewed and approved by the Institutional Review Board at the Icahn School of Medicine at Mount Sinai.

## Results

We identified 29,847 OHCA claims associated with beneficiaries who survived to discharge following OHCA, with mean beneficiary age of 75 years (SD 8); 40.1% female, 80% White; 15.2% Black; 5.8% Other (Table 1). 71.8% (n = 21, 417) of the overall cohort had a significant comorbidity burden (>5) but comorbidity burden was similar between Black and White beneficiaries. Beneficiary level median hospital length of stay (LOS) was 5 days (IQR 0-11 days) and overall, 24.4% (n = 7293) underwent cardiac catheterization and 9.0% (n = 2688) received an ICD during the first admission. The median time-to-event was 533 days (IQR 11.00, 1439.00). Crude survival for the cohort was 54% (n=16,129) at 1 year and 40.8% (n= 12,189) at 3 years.

**Table 1:** Medicare Beneficiary OHCA Cohort Characteristics 2013-2015

| Variable | Overall n (%) | White n (%) | Black n (%) | Other** n (%) |
| --- | --- | --- | --- | --- |
| **Overall OHCA Cohort Beneficiary Level** | 29847 | 23578 (79.0) | 4549 (15.2) | 1720 (5.8) |
| **Sex** |  |  |  |  |
| Female | 11978 (40.1) | 8844 (73.8) | 2464 (20.5) | 670 (5.5) |
| Male | 17869 (59.9) | 14734 (82.4) | 2085 (11.7) | 1050 (5.8) |
| **Age Group Category** |  |  |  |  |
| 65-74 y. | 15131 (50.7) | 11668 (77.1) | 2515 (16.6) | 948 (6.3) |
| 75-84 y. | 10119 (33.9) | 8251 (81.5) | 1385 (13.7) | 483 (4.8) |
| 85+ y. | 4597 (15.4) | 3659 (79.6) | 649 (14.1) | 289 (6.3) |
| **AHRQ Comorbidity Index** |  |  |  |  |
| <0 | 2995 (10.0) | 2482 (10.5) | 366 (8.0) | 147 (8.5) |
| 0 | 2470 (8.3) | 2160 (9.2) | 190 (4.2) | 120 (7.0) |
| 1-4 | 1804 (6.0) | 1544 (6.5) | 162 (3.6) | 98 (5.7) |
| >5 | 21417 (71.8) | 16537 (70.1) | 3586 (78.8) | 1294 (75.2) |
| NA | 1161 (3.9) | 855 (3.6) | 245 (5.4) | 61 (3.5) |
| **Hospital Length of Stay in Days (Median, IQR)** | 5.00 (0.00, 11.00) | 4.00 (0.00, 11.00) | 6.00 (1.00, 14.00) | 7.00 (1.00, 15.00) |
| **Cardiac Catheterization at Index Hospitalization (Y or N)** |  |  |  |  |
| Cardiac Cath Y | 7293 (24.4) | 6110 (25.9) | 753 (16.6) | 430 (25.0) |
| Cardiac Cath N | 22554 (75.6) | 17468 (74.1) | 3796 (83.4) | 1290 (75.0) |
| **Implantable Cardioverter Defibrillator (ICD) at Index Hospitalization (Y or N)** |  |  |  |  |
| ICD Y | 2688 (9.0) | 2241 (9.5) | 301 (6.6) | 146 (8.5) |
| ICD N | 27159 (91.0) | 21337 (90.5) | 4248 (93.4) | 1574 (91.5) |
| Variable | Overall n (%) | White n (%) | Black n (%) | Other n (%) |
| **Index Treating Hospital Characteristics (Beneficiary Level)** |  |  |  |  |
| **Hospital No. of Beds** |  |  |  |  |
| <100 | 4513 (15.1) | 3977 (16.9) | 375 (8.2) | 161 (9.4) |
| 100-399 | 9394 (31.5) | 6901 (29.3) | 1959 (43.1) | 534 (31.0) |
| >400 | 15940 (53.4) | 12700 (53.9) | 2215 (48.7) | 1025 (59.6) |
| NA |  |  |  |  |
| **Hospital Teaching Status** |  |  |  |  |
| Major Teaching | 4165 (14.0) | 2806 (11.9) | 1094 (24.0) | 265 (15.4) |
| Minor Teaching | 10469 (35.1) | 8185 (34.7) | 1623 (35.7) | 661 (38.4) |
| Non-Teaching | 15213 (51.0) | 12587 (53.4) | 1832 (40.3) | 794 (46.2) |
| **Hospital Ownership Status** |  |  |  |  |
| For Profit | 4760 (15.9) | 3669 (15.6) | 773 (17.0) | 318 (18.5) |
| Government (Non-Federal) | 3833 (12.8) | 2915 (12.4) | 685 (15.1) | 233 (13.5) |
| Non-profit | 21254 (71.2) | 16994 (72.1) | 3091 (67.9) | 1169 (68.0) |
| **Medical Intensive Care Unit (MICU) Beds (Y or N)** |  |  |  |  |
| MICU Beds Y | 1948 (6.5) | 1674 (7.1) | 187 (4.1) | 87 (5.1) |
| MICU Beds N | 24013 (80.5) | 18928 (80.3) | 3739 (82.2) | 1346 (78.3) |
| NA | 3886 (13.0) | 2976 (12.6) | 623 (13.7) | 287 (16.7) |
| **Cardiac Intensive Care Beds (CIC) (Y or N)** |  |  |  |  |
| CIC Beds Y | 11607 (38.9) | 9552 (40.5) | 1455 (32.0) | 600 (34.9) |
| CIC Beds N | 14354 (48.1) | 11050 (46.9) | 2471 (54.3) | 833 (48.4) |
| NA | 3886 (13.0) | 2976 (12.6) | 623 (13.7) | 287 (16.7) |
| **Cardiac Catheterization Capability (Y or N)** |  |  |  |  |
| Cardiac Catheterization Y | 20262 (67.9) | 15882 (67.4) | 3193 (70.2) | 1187 (69.0) |
| Cardiac Catheterization N | 9585 (32.1) | 7696 (32.6) | 1356 (29.8) | 533 (31.0) |
| NA |  |  |  |  |
| Outcome | Overall n (%) | Black n (%) | White n (%) | Other n (%) |
| **Alive at 1 year** | 16129 (54.0) | 13187 (55.9) | 1978 (43.5) | 964 (56.0) |
| **Expired at 3 years** | 13718 (46.0) | 10391 (44.1) | 2571 (56.5) | 756 (44.0) |
| **Alive at 3 years** | 12189 (40.8) | 10045 (42.6) | 1392 (30.6) | 752 (43.7) |
| **Expired at 3 years** | 17658 (59.2) | 13533 (57.4) | 3157 (69.4) | 968 (56.3) |

**\*\*Other (CMS Defined Racial and Ethnic Categories):** Hispanic; Asian/Native Hawaiian, or Pacific Islander; American Indian or Alaska Native

The overall survival rates for each quintile by Kaplan-Meier estimation at 1 year for the ICE race measure ranged from Q5 (more segregated White) 54.6% (CI 0.53-0.44) to Q1 (more segregated Black) 46.3% (CI 0.45-0.48) and demonstrated similar rates across the income and racialized measures. For survival at 3 years for the ICE race measure ranged from Q5 42.5% (CI 0.41-0.44) to Q1 32.9% (CI 0.32-0.34); ICE income measure Q5 (high income) 48.3% (CI 0.47-0.50) to Q1 (low income) 32.4% (CI 0.31-0.34); and ICE race + income measure Q5 (high-income White) 48.5% (CI 0.47-0.50) to Q1 (low-income Black) 32% (0.21-0.33) (**see Figure 1)**.

**Figure 1:**
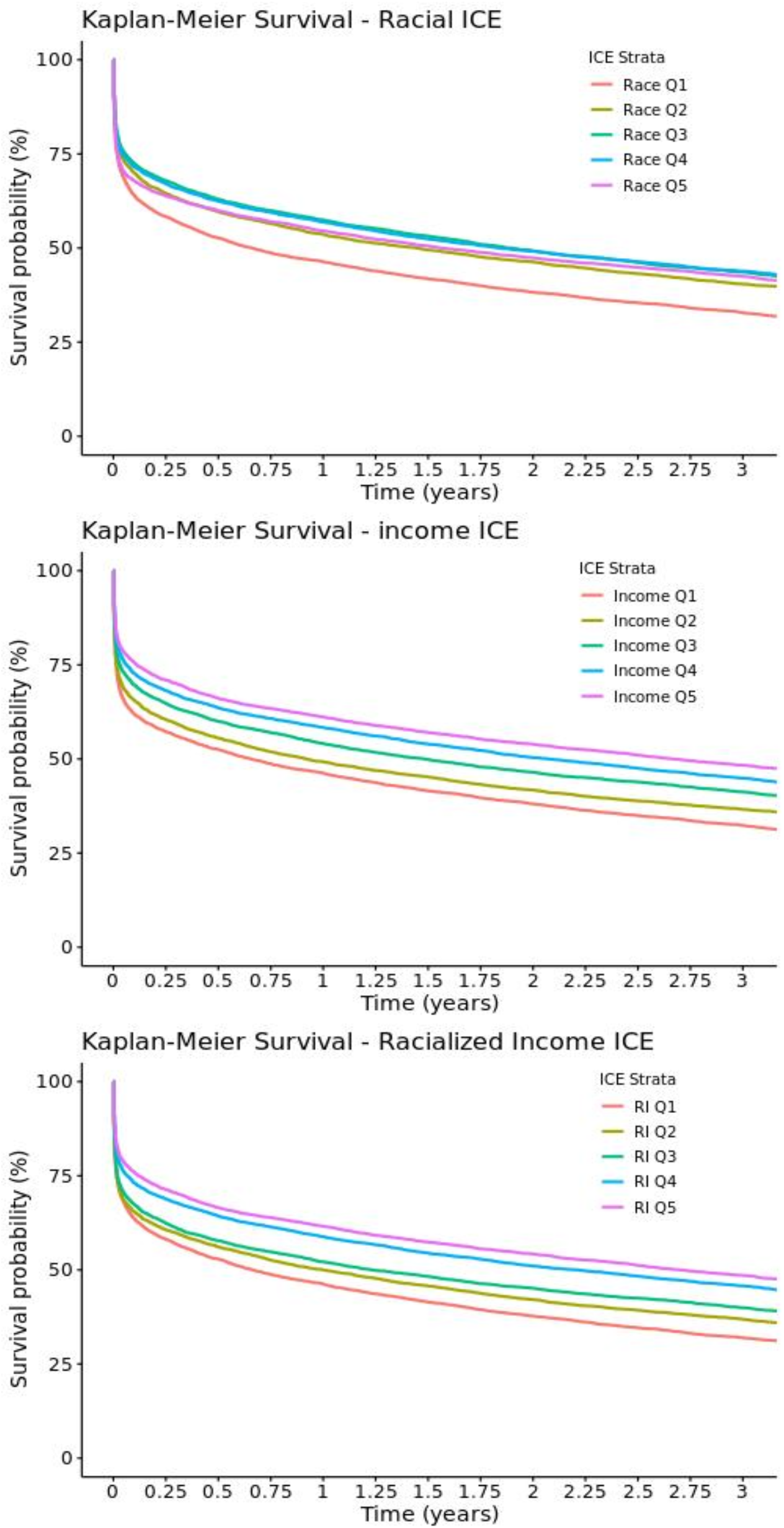
KM Survival Curves for ICE Measures Figure 1A: Kaplan Meier Survival Curve for ICE Measure by Quintile. ICE Race Model Q1 represents most segregated Black and Q5 represents most segregated White. ICE Income Model Q1 represents low income (≤$25,000) and Q5 represents high income (≥$100,000). ICE Income + Race Measure Model Q1 represents low-income Black and Q5 represents high income White.

We found a decreased hazard of death in beneficiaries residing in the most racially and economically privileged ZIP codes (Q5) compared to the least privileged areas (Q1) across all three ICE measures (race: HR:0.84; CI 0.79-0.88, income: HR 0.76; CI 0.73-0.81, race + income: HR 0.78; CI 0.74-0.83) (**Figure 2**). This result was persistent and statistically significant across all ICE measures and quintiles except race + income measure Q2 (HR: 0.99; CI 0.94-1.04). Among beneficiary level covariates in the ICE income and race + income models, Black race was associated with an increased hazard of death (income: HR 1.19; CI 1.14-1.24, race + income: 1.17; CI 1.12-1.23) compared to White beneficiaries, but not observed in the ICE race model. For beneficiaries who underwent interhospital transfer after index OHCA, there was a significant decreased hazard of death across all three measures (HR 0.62; CI 0.59-0.65) compared to beneficiaries not transferred. Among important predictors, beneficiaries not receiving cardiac catheterization and ICD placement at the first treating hospital was strongly associated with increased hazard of death (HR 2.25; CI 2.16-2.35) and (HR 1.95; CI 1.8-2.1), respectively and was seen in all three models as compared to beneficiaries who did (**Supplemental Figures 1-3)**.

**Figure 2:**
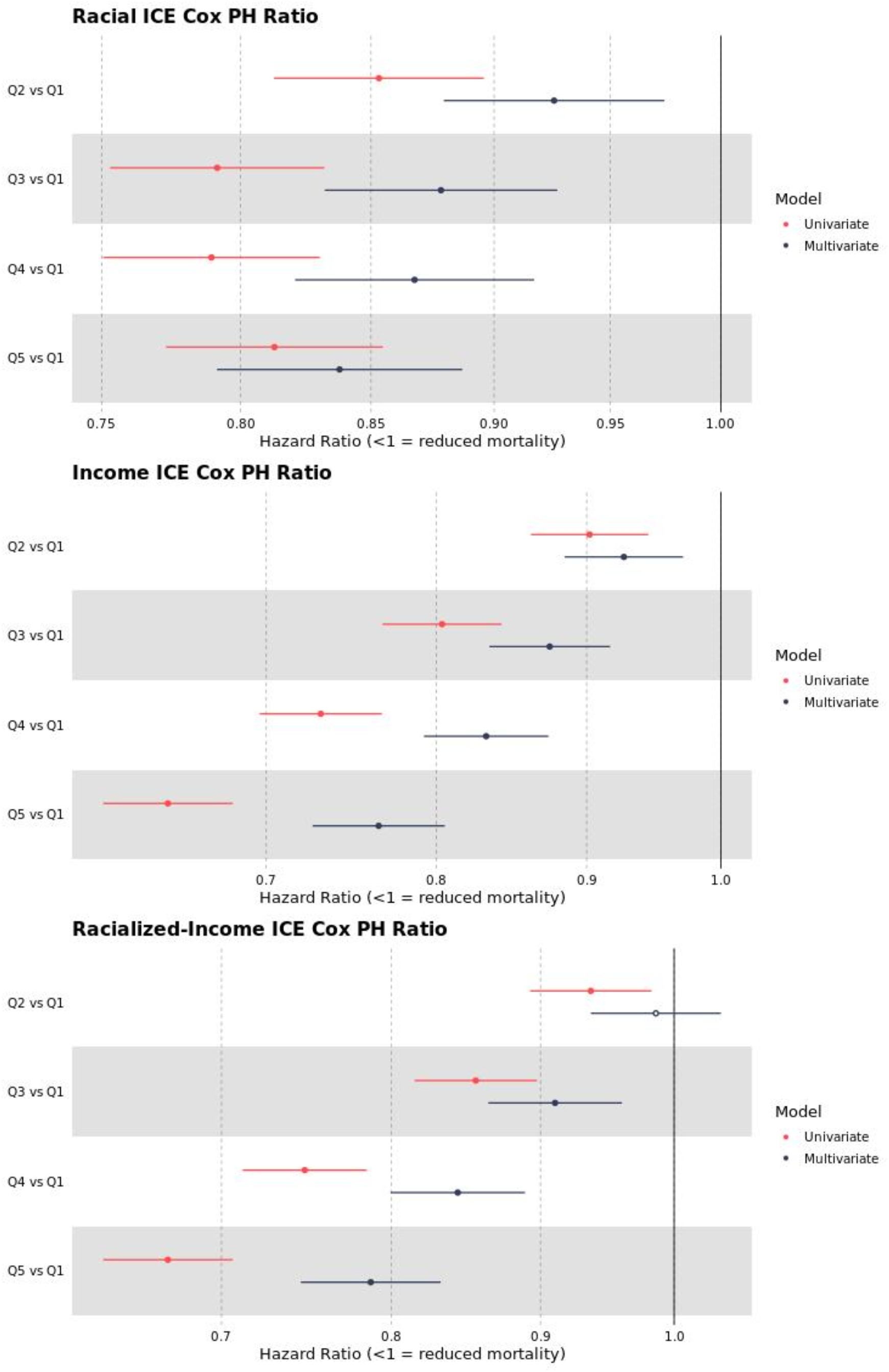
Univariate and Multivariate ICE Cox Proportional Hazards Model Forest Plots Figure 2: Forest Plots of Cox PH Ratios for Univariate and Multivariable Models all ICE Measures. For **Race Measure** Q1=More Segregated Black (ref), Q5=More segregated White. For **Income Measure** Q1 = low income(ref), Q5 =high income. For **Race + Income (Racialized Income)** ICE Measure Q1=More Segregated Black and low income (ref), Q5=More segregated White and high income.

## Discussion

In this study of Medicare beneficiaries who survived to discharge after an OHCA, we demonstrated that among ICE measures of racial and economic segregation there was a decreased hazard of death for those residing in more highly segregated White and higher income ZIP codes relative to those in more highly segregated Black and lower income ZIP codes. For the three ICE measures, the income measure was the strongest predictor, with a 24% decreased hazard of death for those beneficiaries residing in Q5 compared to other quintiles and measures. This finding was present and persistent across all three ICE measures and quintiles except for racialized income measure Q2. Importantly, we also found undergoing ICD placement or cardiac catheterization during index hospitalization was a strong predictor for survival in the fully adjusted models of all three measures.

Our finding of racial disparities in long-term survival after OHCA in the context of validated measures of racial and economic residential segregation contrasts with some previous studies in the cardiac arrest literature. While there is overall limited work examining longitudinal trajectories in OHCA and none known utilizing ICE measures, prior studies have produced mixed results regarding racial and ethnic disparities in long-term survival. Chan and colleagues found 1-year mortality rates of 31.8% and 3-year rates of 47.2% but found no survival differences by individual race when examined in a subgroup analysis in a study utilizing OHCA registry data linked to Medicare claims. Coppler et al. also noted no long-term differences in survival based on race, but found disparities based on sex, socioeconomic status, and geographic post-hospital access to care. This contrasts to a recent in-hospital cardiac arrest study that found lower survival among Black patients at 1, 3, and 5 years after adjustment for hospital site.^31^ Our results suggest that there exists a strong association between racial and economic segregation and long-term survival after OHCA which may be related to important factors related to residential communities and social context post-discharge.

We also found ICD placement and cardiac catheterization were strong predictors for survival across all three ICE measures; beneficiaries who did not undergo those procedures demonstrated an increased hazard for death. Because claims data do not include granular clinical details, we were unable to determine if specific beneficiaries met precise criteria for these interventions. However, our results are in line with several prior studies. A study of Medicare beneficiaries by Groeneveld et al. found a survival benefit for ICD implantation after OHCA for both Black and White beneficiaries. They found that Black beneficiaries had a lower probability of receiving an ICD or coronary revascularization at 90 days among those aged 66-74 years old even after full adjustment which included socioeconomic differences.^32^ Other studies of ICD placement using CMS data found lower utilization of the procedure among Black beneficiaries compared to White beneficiaries but somewhat equalizing by the early 2000’s.^33^ In a contemporary study of elderly in-hospital cardiac arrest survivors, Chen and colleagues found Black patient less likely to undergo coronary revascularization but were as likely to receive an ICD as compared to White survivors.^31^ Our study confirms the survival benefits conferred by ICD placement and cardiac catheterization for beneficiaries after index hospitalization in multivariable models with measures of residential segregation as the primary predictor. Future work will seek if these differences in rates of procedures lead to disparities in outcomes when accounting for individual level race, ethnicity, and income.

While this study was not designed to ascertain causes, racial and economic segregation likely has complex effects on long-term OHCA survival in the context of access to care, quality of care, as well as other important social determinants. Given high rates of readmissions and post-discharge healthcare utilization after an index OHCA,^11, 12^ the structural disinvestment that has been noted to occur in the setting of residential segregation could create barriers to fundamental needs such as transportation, access to follow-up medical care, medications, and multiple other factors important for elderly OHCA survivors. These community factors may play a more important role in longitudinal outcomes however further study is warranted to identify these pathways leading to disparities in outcomes.

Overall, our work shows that where a person lives is independently and significantly associated with their hazard of death after in the three years after discharge for OHCA. This association persists after controlling for patient-level, hospital-level, and hospitalization-specific factors which are independently associated with post-discharge mortality.

### Limitations

There are several noted limitations to our study. Claims data lack the granularity of registry data, and thus we were unable to include important clinical predictors such as initial arrest rhythm, EMS response times, location of arrest, or if bystander CPR was performed in our models. We also note that Medicare data include a sample population that is predominately White, thus limiting our ability to rigorously analyze associations for all racial and ethnic groups in the US. Despite these limitations, we believe that this study captures important population level data on OHCA long-term outcomes among an elderly population. Further, this work closely examines the important intersection and impact of long-standing residential segregation in the US and its association with healthcare outcomes using validated measures.

## Conclusion

In this study of Medicare beneficiaries who survived to discharge after an OHCA, we found a decreased hazard of death for those residing in predominately White and higher income ZIP codes as compared to Black and lower income ZIP codes when using validated measures of racial and economic segregation. Measures of economic segregation based on income were the strongest predictor for long-term survival. This study demonstrates that racial and economic segregation are associated with disparities in outcomes for OHCA. Further study is needed to focus efforts on identifying methods to overcome systemic and institutional racism to achieve equity in health outcomes for OHCA and other cardiovascular conditions.

## Data Availability

All data produced in the present work are contained in the manuscript. CMS data use agreements preclude any sharing of data with third parties.

---

**Figure 1:** Kaplan Meier Survival Curve for ICE Race, Income, and Race + Income Models

**Figure 2:** Univariate and Multivariate ICE Measures Cox Proportional Hazards Model Forest Plot

**Supplementary Figure 1:** Multivariate ICE Race Cox Proportional Hazards Model Forest Plot (all covariates)

**Supplementary Figure 2:** Multivariate ICE Income Cox Proportional Hazards Model Forest Plot (all covariates)

**Supplementary Figure 3:** Multivariate ICE Race + Income Cox Proportional Hazards Model Forest Plot (all covariates)

**Supplementary Figure 1:**
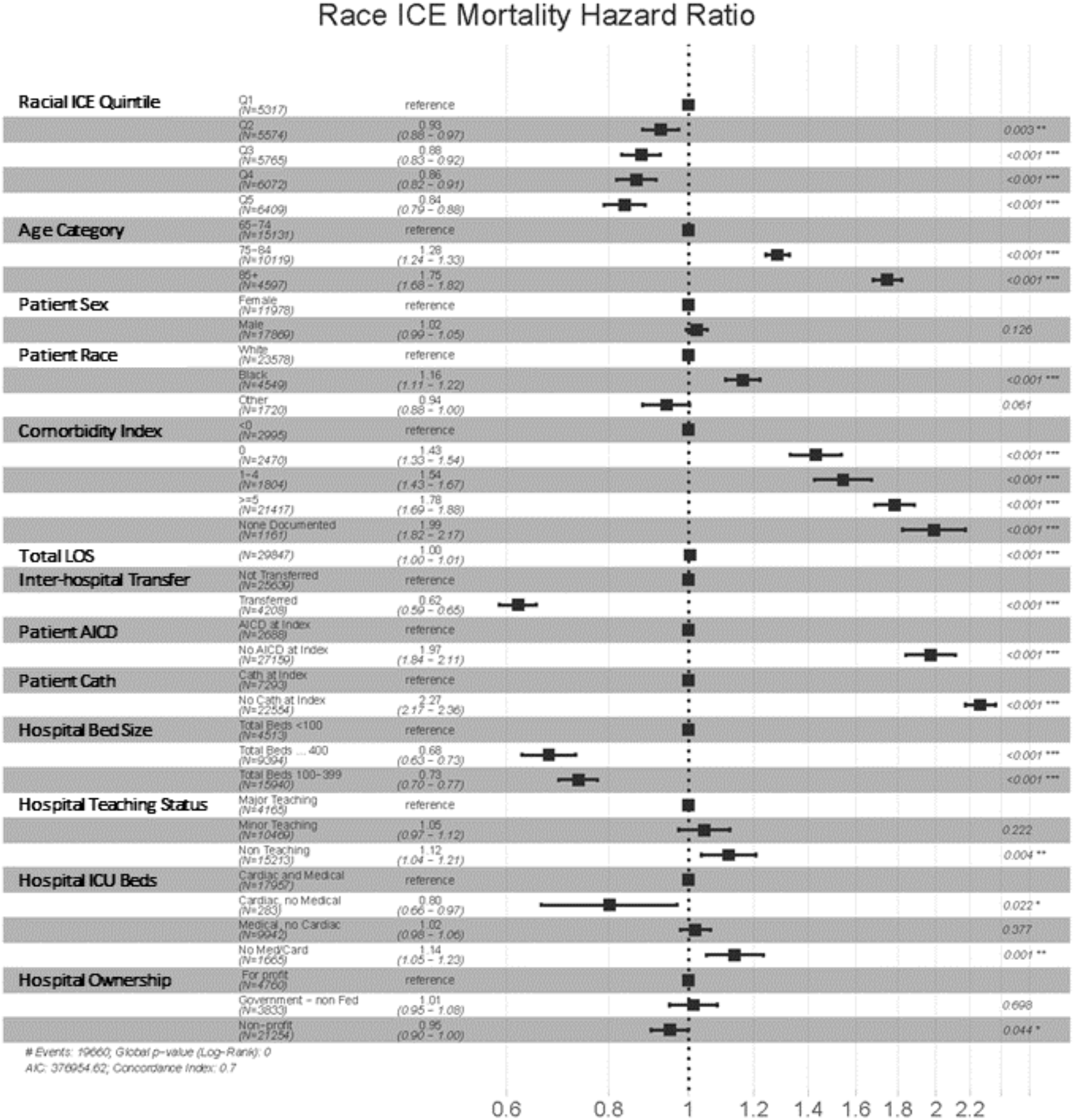
Race ICE Measure Cox PH Multivariable Model Hazard Ratios and 95% Confidence Intervals

**Supplementary Figure 2:**
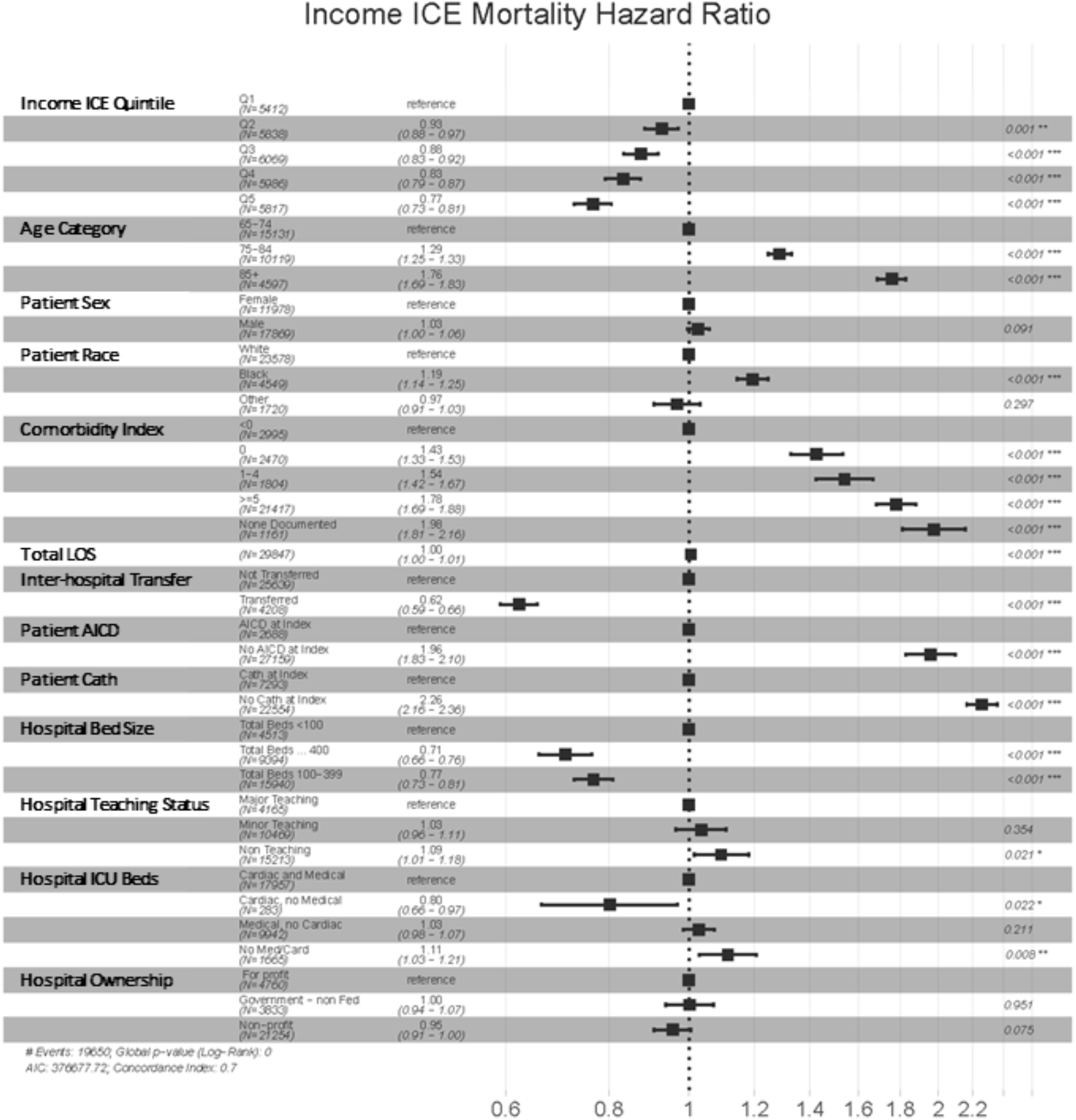
Income ICE Measure Cox PH Multivariable Model Hazard Ratios and 95% Confidence Intervals

**Supplementary Figure 3:**
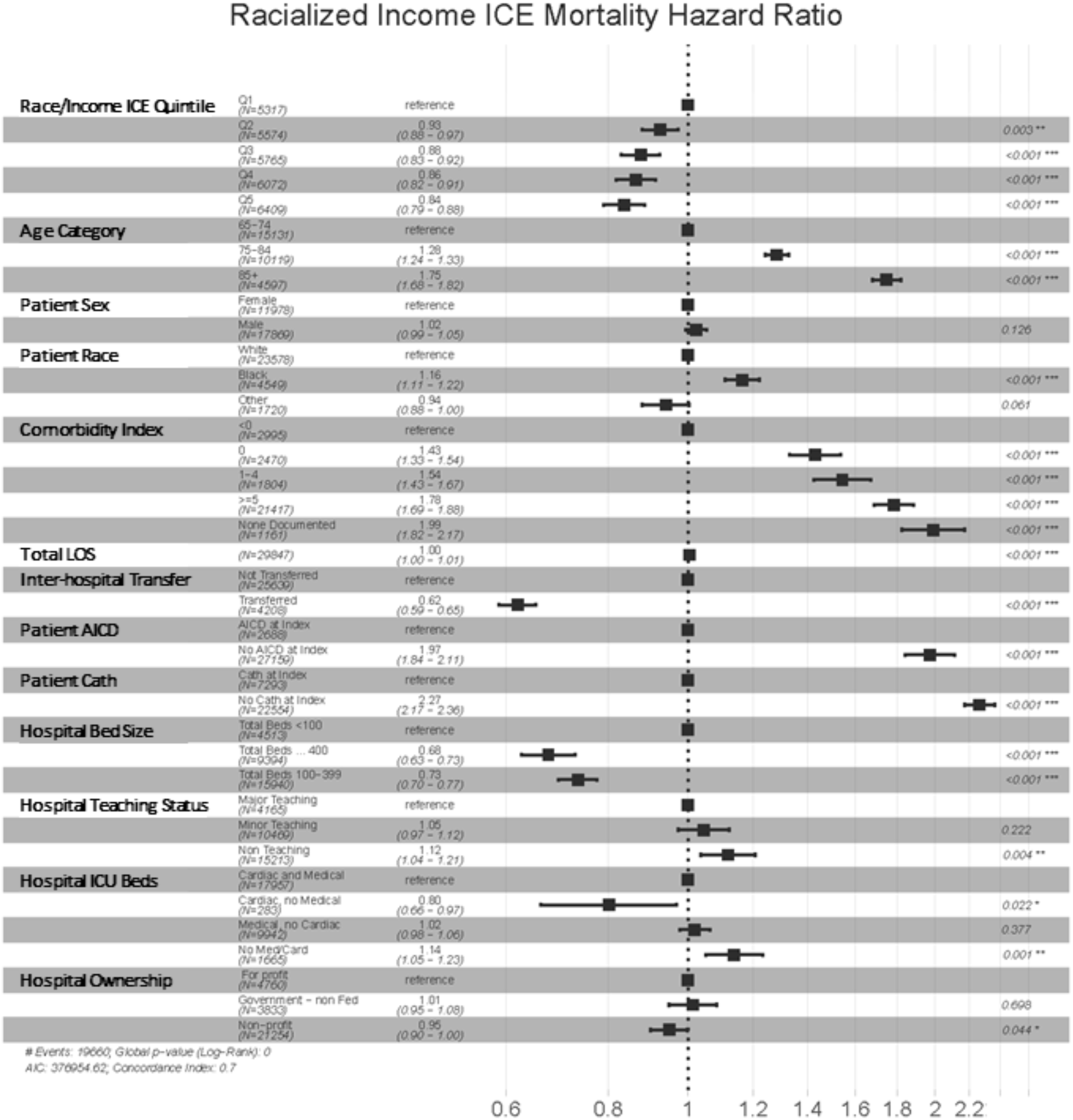
Race + Income (racialized income) ICE Measure Cox PH Multivariable Model Hazard Ratios and 95% Confidence Intervals

